# Prevalence of Urogenital Schistosomiasis and Associated Factors in the Lac Region: A Focus on Environmental and Socioeconomic Contexts

**DOI:** 10.64898/2026.07.26.26358946

**Authors:** Didier Lalaye, Marius Madjissem, Loukoum Nan-Arabé

**Affiliations:** Utrecht Medical Centre, Utrecht University; MD Bol District Hospital; Faculty of Human Health Sciences, University of N’Djamena

**Keywords:** Schistosoma haematobium, urogenital schistosomiasis, prevalence, Lake Chad, Chad, mobile health, logistic regression

## Abstract

**Background:** Urogenital schistosomiasis caused by Schistosoma haematobium remains a major public health concern in the Lake Chad region. This study aimed to estimate the cumulative prevalence of S. haematobium infection and to identify factors associated with test positivity within an intervention zone of the Ngouri health district, Lac Province, Chad.

**Methods:** A cross-sectional analytical study was conducted using routinely collected data from the Dawa Mobile Health system between February 2024 and April 2025. A total of 4,504 individuals were included after data cleaning and biological consistency correction (22 haematuria cases recoded as positive). Bivariate analysis (Chi-square test) and multivariable logistic regression were performed.

**Results:** The overall prevalence was 19.5% (95% CI: 18.4%–20.7%). No statistically significant association was found for age (p=0.163), sex (p=0.939), marital status (p=0.858), occupation (p=0.704), or urine appearance in the binary model (p=0.116). Education level approached significance (p=0.078), with the highest prevalence observed at the primary level (23.0%). In multivariable analysis, only primary education was independently associated with positivity (adjusted OR=1.29, 95% CI: 1.06–1.57; p=0.011).

**Conclusion:** Nearly one in five participants tested positive, confirming the high endemicity of the Ngouri district. The homogeneous distribution across demographic subgroups suggests widespread community exposure, supporting mass drug administration beyond school-aged children, strengthened WASH interventions, and consolidation of mobile health surveillance.

## INTRODUCTION

Urogenital schistosomiasis, caused by Schistosoma haematobium, remains one of the most prevalent parasitic diseases in sub-Saharan Africa, constituting a major public health burden in communities whose daily activities necessitate contact with freshwater bodies [1, 2]. Transmission occurs through exposure to water contaminated with cercariae released by infected intermediate host snails of the genus Bulinus. Because exposure is intimately linked to living conditions and water-use behaviors, infection tends to cluster within specific ecological and social contexts, disproportionately affecting populations with frequent water contact — notably school-aged children, young adults, and individuals engaged in farming, fishing, irrigation, or other water-related occupations [3, 4].

Recent environmental changes, particularly those driven by climate variability and rising temperatures, have further exacerbated the challenges faced by these communities. Shifting rainfall patterns and increasing temperatures have altered water availability and expanded stagnant water bodies, thereby creating favorable habitats for intermediate host snails [5]. In the Lake Chad region, the progressive shrinkage of the lake — one of the most dramatic hydrological changes observed in Africa over recent decades — has paradoxically given rise to the emergence and persistence of small, stagnant water pockets that serve as critical water sources for livestock and agriculture [6]. These residual water bodies may harbor infective cercariae, placing both animals and their caretakers at heightened risk. Children who herd livestock or play near water sources, as well as women engaged in water collection, washing, and agricultural activities, are particularly vulnerable to repeated exposure [7].

Beyond its direct morbidity, S. haematobium infection carries substantial clinical and socioeconomic consequences. Chronic urogenital schistosomiasis may lead to hematuria, lower urinary tract symptoms, bladder wall pathology, obstructive uropathy, and long-term sequelae including bladder cancer and, in women, female genital schistosomiasis associated with increased susceptibility to HIV [8, 9]. These outcomes can impair school attendance, reduce economic productivity, and intensify the burden on already fragile health systems. Compounding this challenge, many infections remain undiagnosed due to the intermittent or nonspecific nature of symptoms and the limited availability of laboratory confirmation in peripheral health facilities [10]. Control strategies therefore rely heavily on prevalence surveys to guide preventive chemotherapy with praziquantel, health education, and water, sanitation, and hygiene (WASH) interventions [1].

However, disease prevalence is not static. It varies according to geography, seasonality, transmission intensity, population mobility, and — critically — the implementation and coverage of control measures [11]. In the Lake Chad region, our initial baseline survey conducted prior to our large-scale intervention activities estimated a local prevalence of 33.1%. Building on this assessment, a 15-month intervention period was subsequently implemented, combining community sensitization with systematic mass drug administration of praziquantel [12].

The prevalence estimate presented in this study therefore represents a cumulative prevalence calculated over the entire duration of the project and across all exhaustively screened participants. It does not reflect the underlying prevalence of urogenital schistosomiasis in the broader Lake Chad region in the absence of intervention. Rather, it captures the epidemiological situation within a defined population exposed to sustained awareness-raising activities and repeated treatment over 15 months. As such, the findings should be interpreted as the prevalence observed within an active intervention zone, and not as a region-wide, community-representative estimate.

Nevertheless, generating updated local data within an intervention context remains epidemiologically essential. Understanding which individual-level characteristics are associated with persistent infection helps refine targeting strategies, optimize resource allocation, and identify subgroups requiring intensified follow-up. Demographic factors such as age and sex often capture differences in behavioral exposure and acquired immunity, while socio-demographic characteristics — including marital status, education level, and occupation — may serve as proxies for exposure patterns and socioeconomic vulnerability. Furthermore, simple clinical indicators such as the macroscopic appearance of urine may provide useful screening clues in resource-constrained settings.

This study therefore aimed to estimate the cumulative prevalence of S. haematobium infection within the Ngouri intervention area in Lac Province, Chad, and to identify factors associated with test positivity, examining demographic, socio-demographic, and clinical variables through bivariate analyses and multivariable logistic regression. The findings are intended to inform ongoing schistosomiasis control efforts and contribute context-specific evidence for programmatic monitoring and strategic planning.

## MATERIALS AND METHODS

### Study Design and Period

This study employed a cross-sectional analytical design with both retrospective and prospective components, based on routinely collected programmatic data from the Ngouri health district, Lac Province, Chad, from February 2024 to April 2025. The study was implemented using the Dawa Mobile Health system, a digital health platform previously demonstrated to be effective in the detection and management of urogenital schistosomiasis in the Torrock area of Chad [13].

A total of 4,570 individuals were initially enrolled through systematic village-level recruitment. Cases with incomplete or inconsistent data were subsequently excluded. A biological consistency correction was applied: 22 observations with haematuria coded as negative were recoded as positive, as macroscopic haematuria is a direct clinical marker of S. haematobium infection. Each participant was assigned a unique identification code recorded in a standardized registry. Sociodemographic and clinical data were collected using a structured questionnaire, and a urine specimen was obtained from each subject.

### Study Area

The Ngouri sub-prefecture is located in Lac Province, western Chad, approximately 230 km north of N’Djamena. The area is predominantly agro-pastoral, covering 3,375 km² with an estimated population of 262,250 inhabitants and a sex ratio of 0.98 (https://fr.city-facts.com/ngouri-td/population). The Ngouri Health District is one of the seven operational districts within the Provincial Health Delegation of Lac. The progressive desiccation of Lake Chad, largely attributed to climate change and demographic pressure, has led to the emergence of residual stagnant water bodies in surrounding villages, creating favorable conditions for Bulinus spp., the intermediate host snails of S. haematobium, thereby sustaining active transmission cycles [14]. The district borders Mao (Kanem) to the northwest, Kouloudia to the south, Mondo and Massakory to the east, and Dibinintchi and Isseirom to the west. It is served by 20 peripheral health centres (most without laboratories) and one district hospital in Ngouri. No prior epidemiological study on urogenital schistosomiasis had been conducted in this locality, and no mass drug administration campaign had been implemented before this project.

### Study Population

The study population comprised all individuals residing in the Ngouri district who were screened for urogenital schistosomiasis through urine-based testing during the study period within the Dawa Mobile Health program.

#### Inclusion criteria

- Permanent residents of Lac Province during the study period
- Individuals who underwent urine testing for S. haematobium within the program
- Availability of complete data on infection status and all selected covariates

#### Exclusion criteria

- Missing or uninterpretable parasitological test result
- Incomplete sociodemographic information precluding variable classification Final sample analysed: **4,569 individuals.**

### Data Collection

Data were collected prospectively through routine screening activities. The following variables were systematically recorded for each participant: age group, sex, marital status, education level, occupation, macroscopic urine appearance, and parasitological test result.

### Diagnostic Procedure

Urine specimens were collected between 9:00 a.m. and 1:00 p.m., corresponding to peak cercarial activity and maximal egg excretion [15]. Children were weighed at the time of collection to enable weight-based praziquantel dosing. All specimens were transported to the district hospital laboratory within three hours of collection.

Samples were first screened for haematuria by dipstick; positive samples were processed by polycarbonate membrane filtration (pore size: 12–20 µm) for microscopic detection and quantification of S. haematobium eggs, in accordance with the standard WHO filtration technique [15]. Infection status was defined as positive when at least one S. haematobium egg was detected per 10 mL of urine. Positive cases received praziquantel at 40 mg/kg body weight free of charge.

The Dawa Mobile Health workflow was as follows: (1) a trained community health agent collected specimens and transported them to the laboratory; (2) the laboratory technician processed samples and notified the physician by SMS; (3) the physician calculated the praziquantel dose and sent the prescription electronically to the pharmacist; (4) the pharmacist prepared labeled medication and coordinated delivery with the field agent by SMS.

### Data Management and Statistical Analysis

Raw data were reviewed for completeness, consistency, and plausibility. A biological consistency correction was applied prior to analysis: 22 observations with haematuria coded as negative were recorded as positive. Variables with non-standard entries or ambiguous categories were harmonized following pre-specified rules.

Overall prevalence was estimated with 95% confidence intervals (CIs) using the Wilson score method. Associations between infection status and explanatory variables were assessed by Pearson’s Chi-square test, with crude odds ratios (ORs) estimated by simple logistic regression (selection threshold p < 0.20 for multivariable inclusion). A multivariable binary logistic regression model was constructed [10] with age and sex included a priori. Urine appearance was recoded as binary (Normal vs. Abnormal = Turbid + Haematic) due to complete separation following the haematuria correction. Model adequacy was assessed using the McFadden pseudo-R² statistic. Statistical significance was set at p < 0.05. All analyses were performed using Python 3.11 (statsmodels 0.14, scipy 1.11).

### Ethical Considerations

Written administrative authorization was obtained from the Ngouri district health authority prior to data collection. Oral informed consent was obtained from all adult participants and from the parents or legal guardians of minor participants. Consent was documented in the enrollment registry. All data were anonymized before analysis. An exemption from full ethical review was granted by the National Bioethics Committee of Chad (Comité National de Bioéthique du Tchad).

## RESULTS

### 1. General Characteristics of the Study Population

A total of 4,504 individuals were retained for analysis after exclusion of records with missing or inconsistent data. The population was predominantly young: the 11–20 year age group was the largest, comprising 1,534 participants (34.1%), followed by the 21–30 year group (1,330 participants, 29.5%). Children aged 1–10 years represented 8.9% of the sample (n = 402).

Females were slightly in the majority (2,298, or 51.0%) compared to 2,206 males (49.0%), with a sex ratio of 1.04. The most frequent marital status was married (3,036 subjects, 67.4%), followed by single (1,402 subjects, 31.1%). The majority of participants had no formal schooling (2,956 subjects, 65.6%), while primary and secondary education accounted for 16.0% and 16.9% of the sample, respectively. The most common occupations were housewife (1,848 subjects, 41.0%), student/pupil (1,162 subjects, 25.8%), and farmer (998 subjects, 22.2%). Regarding macroscopic urine appearance, it was described as normal in 2,886 cases (64.1%), turbid in 1,590 cases (35.3%), and haematic in 28 cases (0.6%).

**Table 1.** General characteristics of the study population (N = 4,504)

| Variable | Category | N | % |
| --- | --- | --- | --- |
| Age | 1–10 years | 402 | 8.9 |
|  | 11–20 years | 1,534 | 34.1 |
|  | 21–30 years | 1,330 | 29.5 |
|  | 31–40 years | 624 | 13.9 |
|  | 41 years and + | 614 | 13.6 |
| Sex | Female | 2,298 | 51.0 |
|  | Male | 2,206 | 49.0 |
| Marital status | Married | 3,036 | 67.4 |
|  | Single | 1,402 | 31.1 |
|  | Widowed | 37 | 0.8 |
|  | Divorced | 29 | 0.6 |
| Education level | No schooling | 2,956 | 65.6 |
|  | Primary | 721 | 16.0 |
|  | Secondary | 763 | 16.9 |
|  | Higher/University | 64 | 1.4 |
| Occupation | Housewife | 1,848 | 41.0 |
|  | Student/Pupil | 1,162 | 25.8 |
|  | Farmer | 998 | 22.2 |
|  | No occupation | 201 | 4.5 |
|  | Teacher/Manager | 106 | 2.4 |
|  | Others | 189 | 4.2 |
| Urine appearance | Normal | 2,886 | 64.1 |
|  | Turbid | 1,590 | 35.3 |
|  | Haematic* | 28 | 0.6 |

\* 22 haematic cases initially coded as "Negative" were recoded as "Positive" (biological consistency correction).

### 2. Overall Prevalence of Urogenital Schistosomiasis

Of the 4,504 individuals analysed, 897 tested positive for Schistosoma haematobium after data correction, yielding an **overall prevalence of 19.5% (95% CI: 18.4%–20.7%)**, estimated using the Wilson score method. Approximately one in five participants had an active infection in this intervention zone. The remaining 3,607 subjects (80.5%) tested negative.

**Figure 1.**
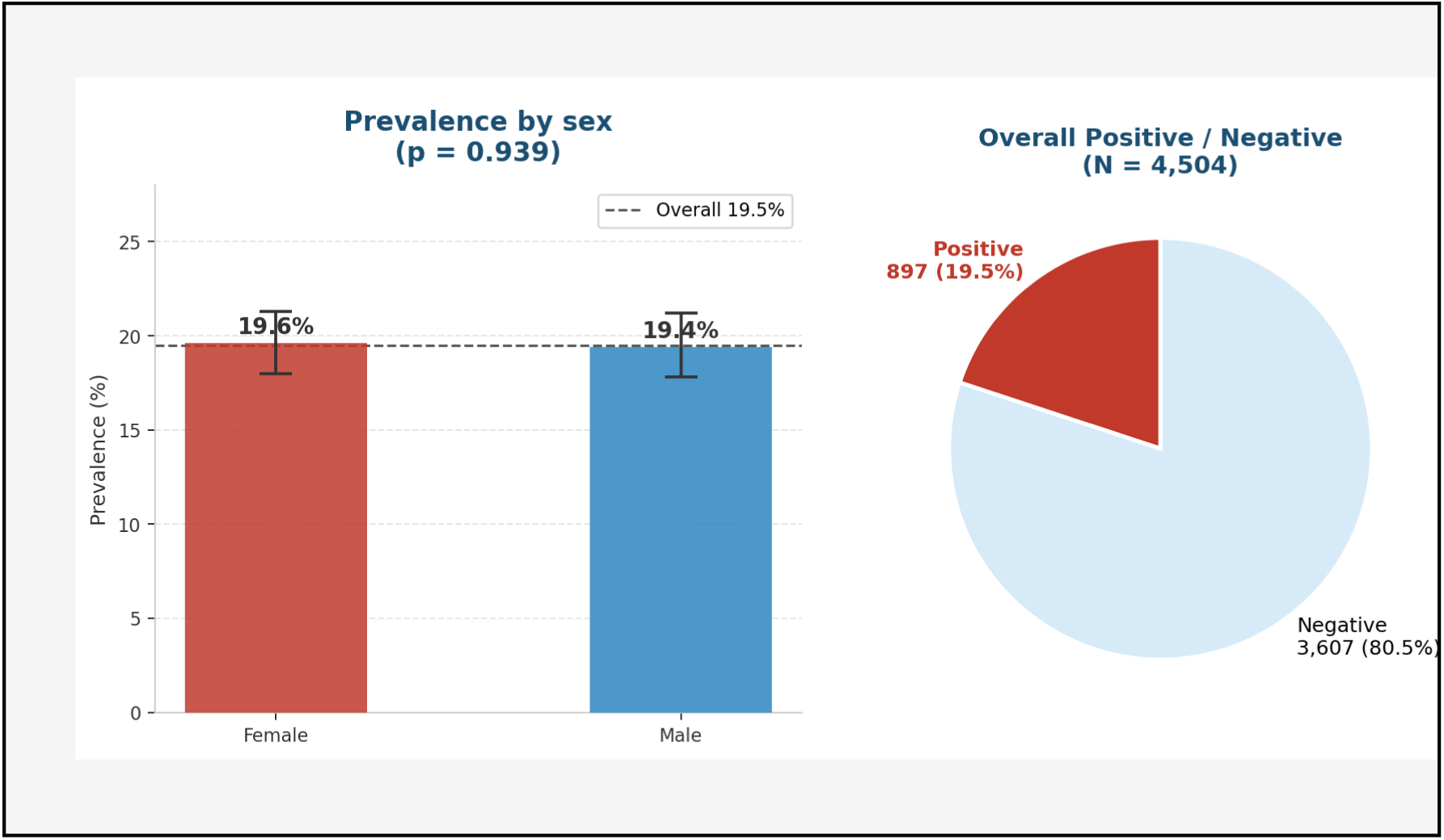
Breakdown of positive and negative cases (pie chart) and prevalence by sex (bar chart). Female: 19.6% vs Male: 19.4% — p = 0.939 (non-significant). Dashed line = overall prevalence (19.5%).

### 3. Prevalence by Age Group and Sex

Prevalence varied moderately across age groups without reaching statistical significance (Chi² = 6.54; df = 4; p = 0.163). The 31–40 year group showed the highest prevalence (21.8%; 95% CI: 18.7%–25.2%), followed by the 11–20 year group (20.4%). Regarding sex, prevalence was virtually identical between females (19.6%; 95% CI: 18.0%–21.3%) and males (19.4%; 95% CI: 17.8%–21.2%), with no statistically significant difference (p = 0.939). The infection was therefore homogeneously distributed across both age groups and sex categories.

**Figure 2.**
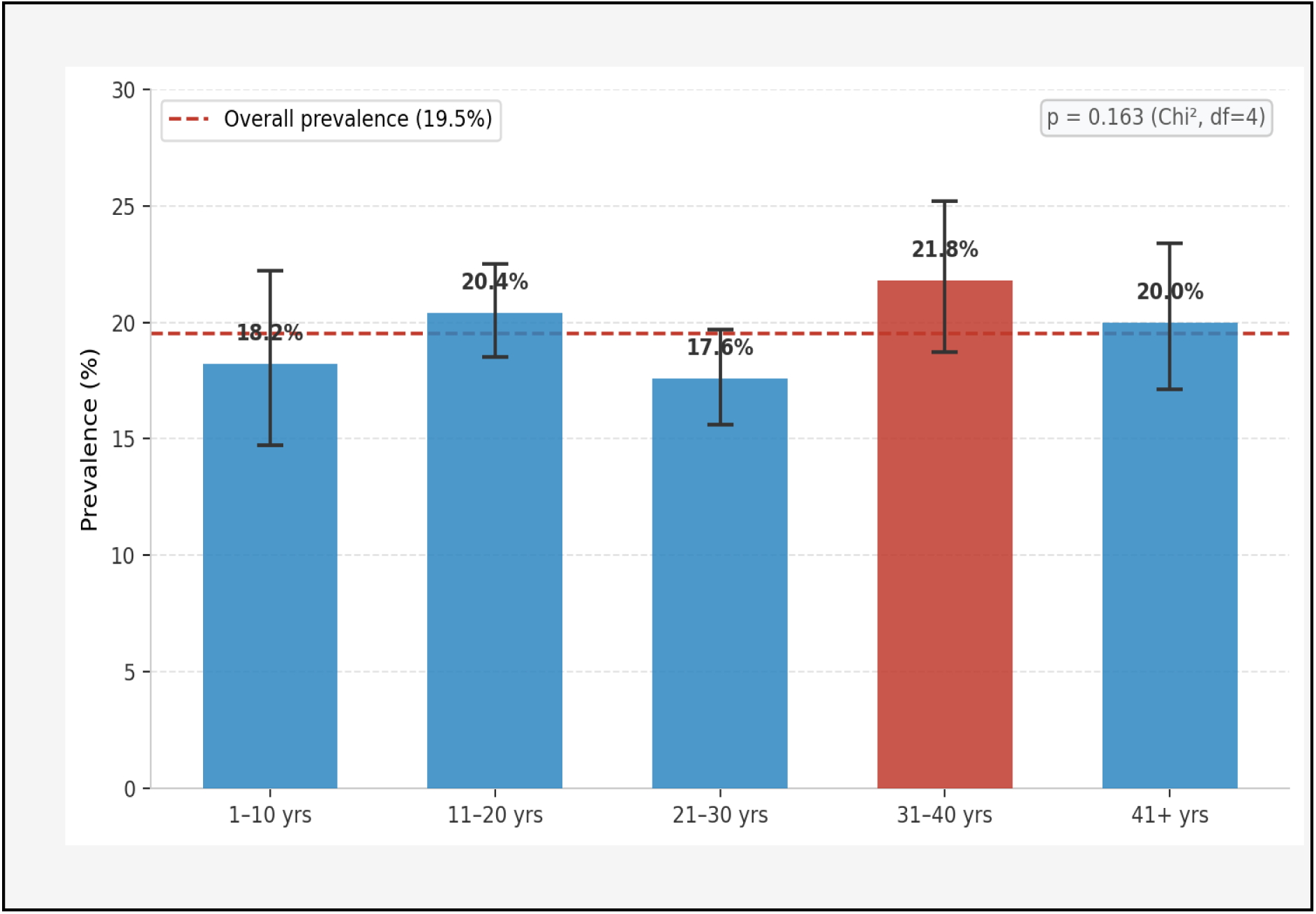
Prevalence of S. haematobium by age group with 95% confidence intervals. The red bar indicates the 31–40 year group (highest prevalence: 21.8%). Dashed line = overall prevalence (19.5%). p = 0.163 (Chi², df = 4).

**Table 2.**
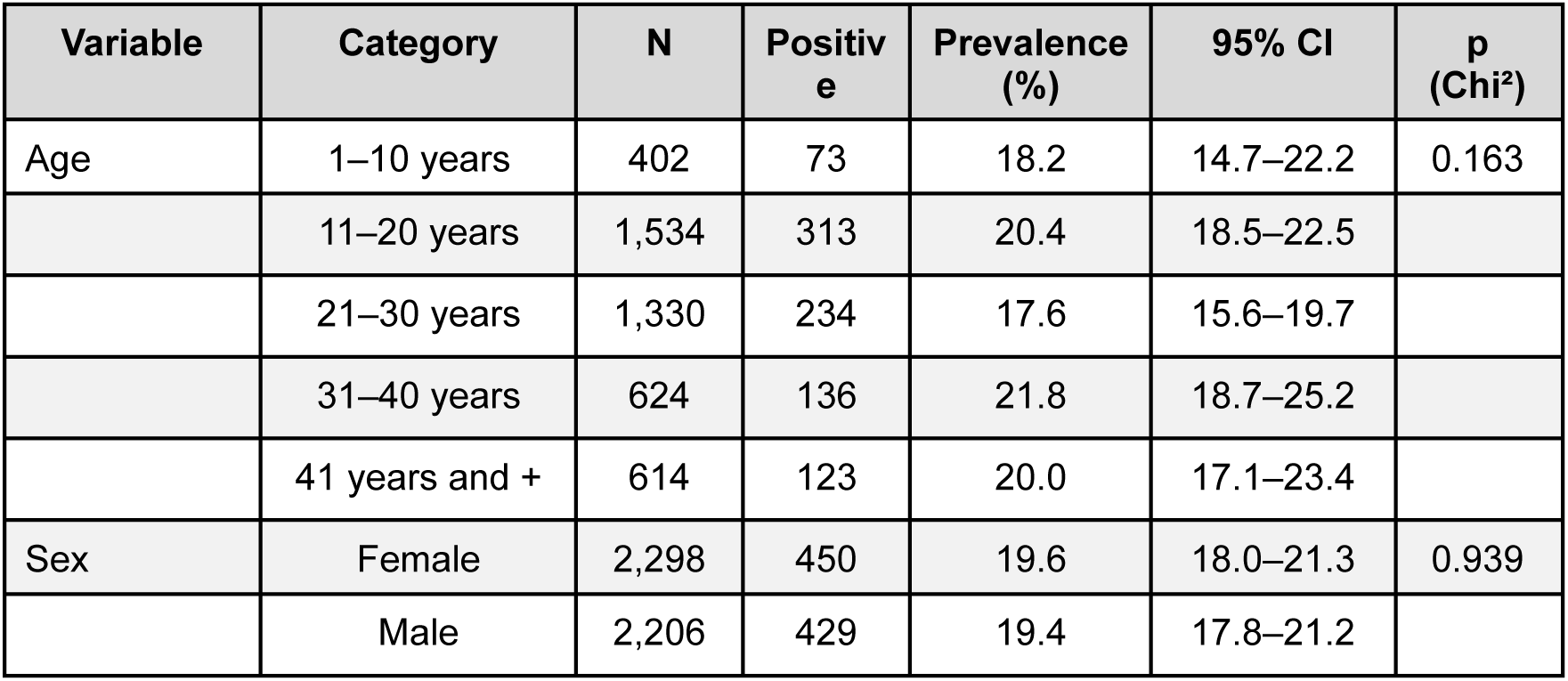
Prevalence by age group and sex.

### 4. Prevalence by Education Level

Education level approached statistical significance (Chi² = 6.81; df = 3; p = 0.078). Participants with primary education had the highest prevalence of all subgroups analysed (23.0%; 95% CI: 20.1%–26.2%), representing an excess of 3.5 percentage points above the overall average. Participants with no schooling and those with secondary education showed similar prevalences (18.8% and 18.9%, respectively).

**Figure 3.**
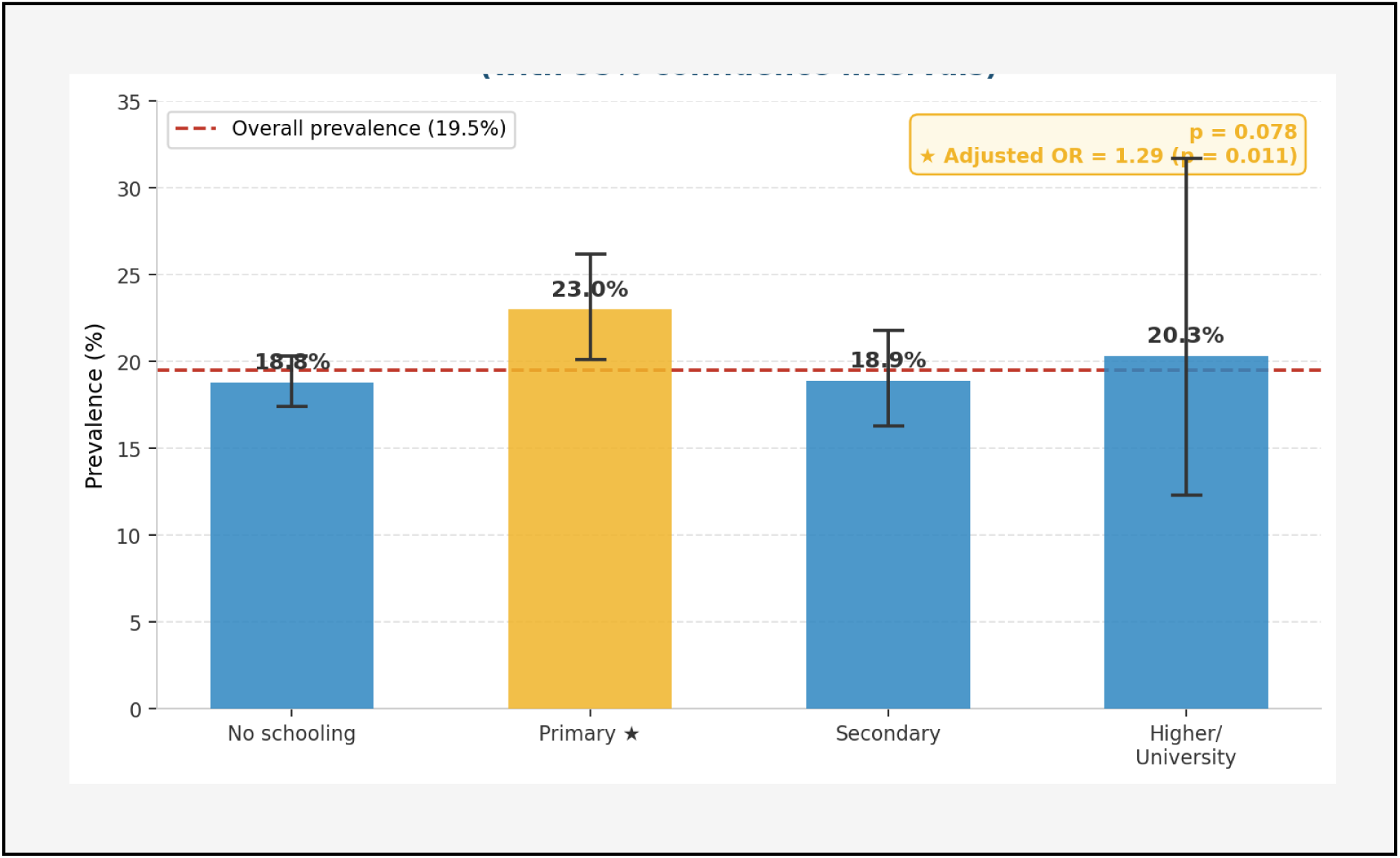
Prevalence of S. haematobium by education level with 95% confidence intervals. The yellow bar indicates primary education (highest prevalence: 23.0%). Dashed line = overall prevalence (19.5%). ★ Only significant factor in multivariable analysis (adjusted OR = 1.29; p = 0.011). Overall p = 0.078.

**Table 3.** Prevalence by education level.

| Education level | N | Positive | Prevalence (%) | 95% CI |
| --- | --- | --- | --- | --- |
| No schooling | 2,956 | 556 | 18.8 | 17.4–20.3 |
| Primary ★ | 721 | 166 | 23.0 | 20.1–26.2 |
| Secondary | 763 | 144 | 18.9 | 16.3–21.8 |
| Higher/University | 64 | 13 | 20.3 | 12.3–31.7 |

★ $p = 0.011$ in multivariable analysis — adjusted OR = 1.29 (95% CI: 1.06–1.57).

### 5. Prevalence by Occupation

No statistically significant association was found between occupational category and test positivity (Chi² = 4.64; df = 7; p = 0.704). Prevalences ranged from 18.4% among housewives to 20.9% among those with no occupation, with no clinically or statistically meaningful difference between groups.

**Figure 4.**
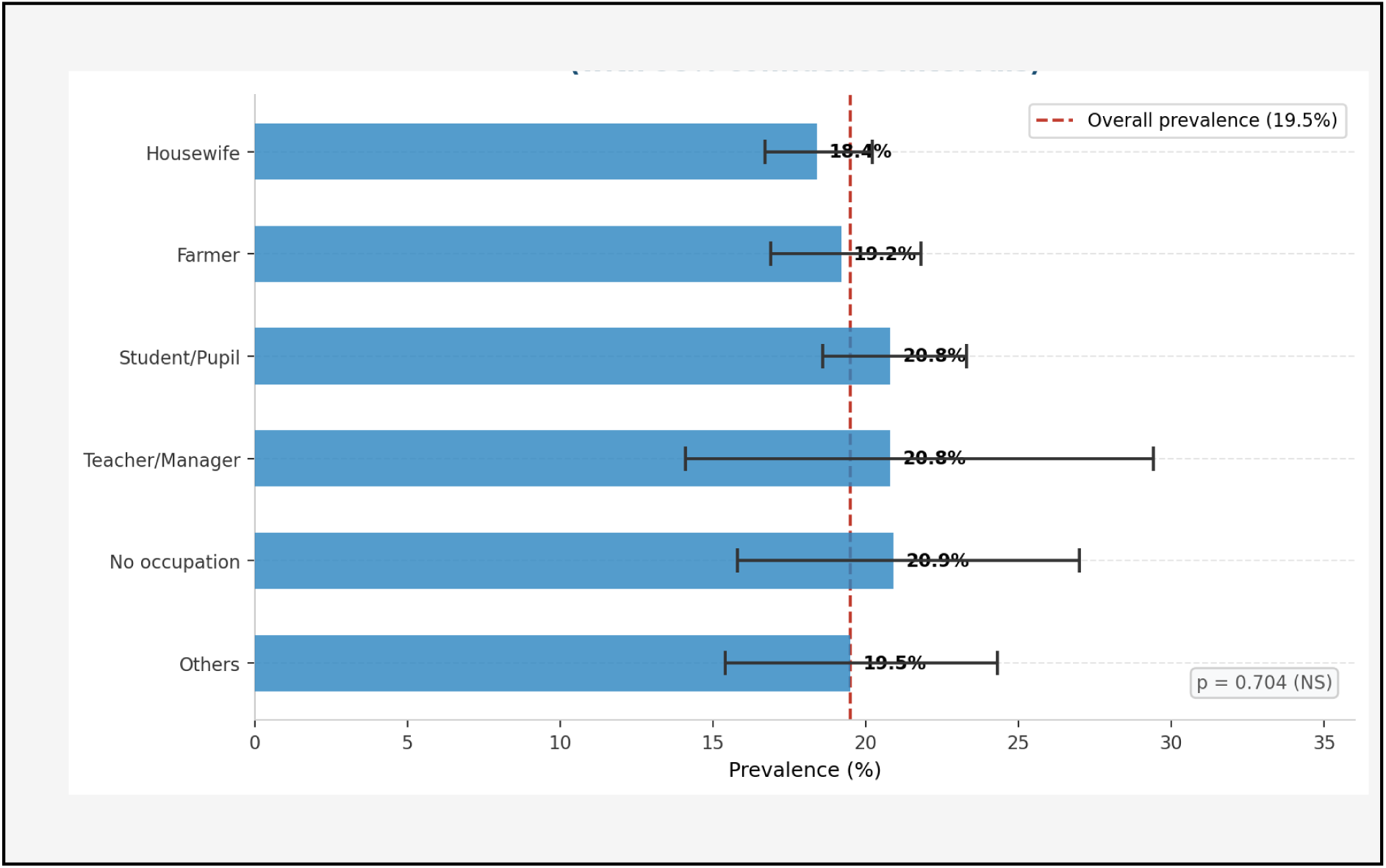
Prevalence of S. haematobium by occupation with 95% confidence intervals (horizontal bars). Dashed line = overall prevalence (19.5%). p = 0.704 (non-significant).

**Table 4.** Prevalence by occupation.

| Occupation | N | Positive | Prevalence (%) | 95% CI |
| --- | --- | --- | --- | --- |
| Housewife | 1,848 | 340 | 18.4 | 16.7–20.2 |
| Student/Pupil | 1,162 | 242 | 20.8 | 18.6–23.3 |
| Farmer | 998 | 192 | 19.2 | 16.9–21.8 |
| Teacher/Manager | 106 | 22 | 20.8 | 14.1–29.4 |
| No occupation | 201 | 42 | 20.9 | 15.8–27.0 |
| Others | 189 | 59 | 19.5 | 15.4–24.3 |

### 6. Multivariable Analysis — Logistic Regression

A binary logistic regression model was fitted including all explanatory variables simultaneously. After adjustment, only one factor remained independently and significantly associated with test positivity: primary education level (adjusted OR = 1.29; 95% CI: 1.06–1.57; p = 0.011), compared to participants with no schooling as the reference. No other variable reached statistical significance after adjustment. The McFadden pseudo-R² was 0.005, reflecting the homogeneous distribution of infection across the study population.

**Figure 6.**
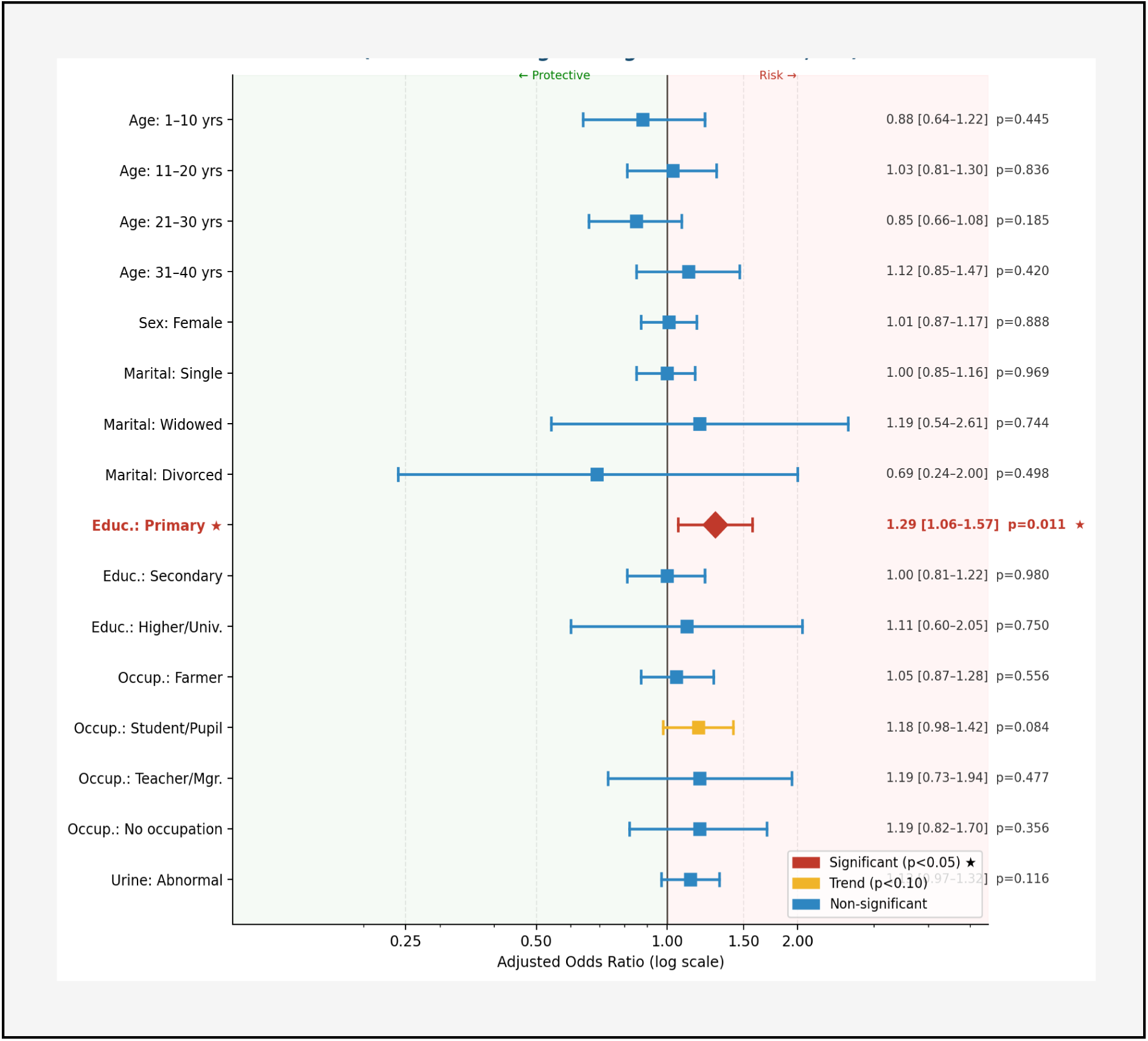
Forest plot of adjusted odds ratios (multivariable logistic regression, N = 4,504). ★ Red: significant (p<0.05). Yellow: trend (p<0.10). Blue: non-significant. Log scale. Vertical line = reference OR (1.0). Only primary education is significant (OR = 1.29; p = 0.011).

**Table 6.**
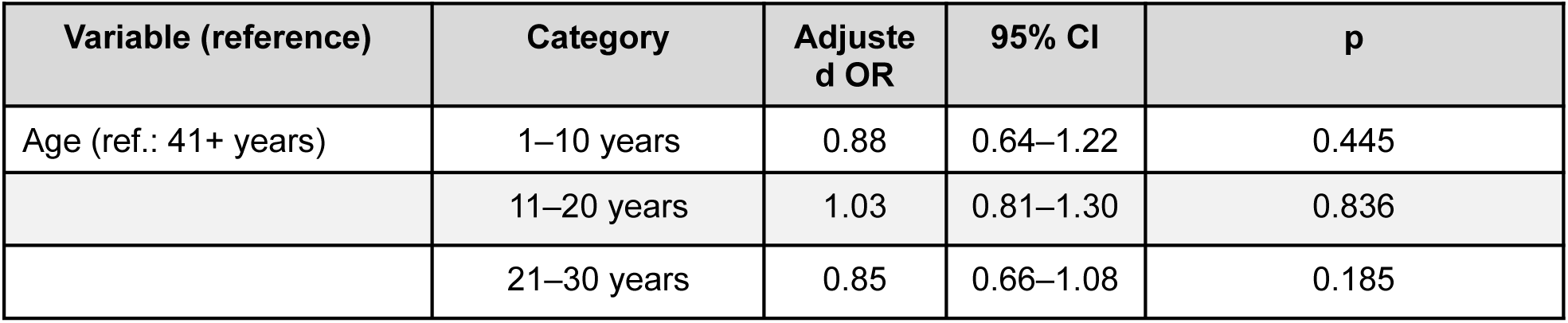
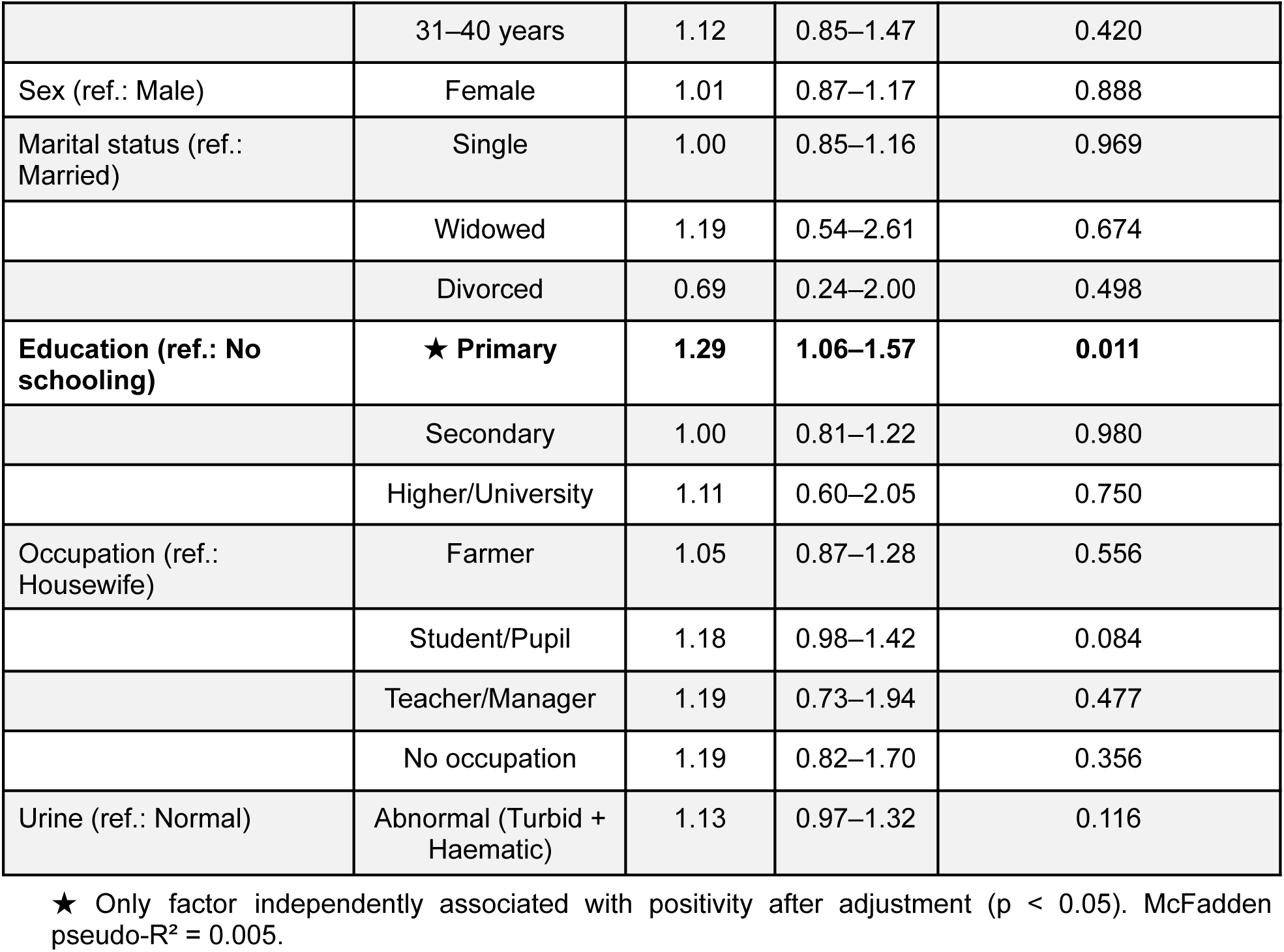
Multivariable logistic regression results (N = 4,504)

| Variable (reference) | Category | Adjusted OR | 95% CI | p |
| --- | --- | --- | --- | --- |
| Age (ref.: 41+ years) | 1–10 years | 0.88 | 0.64–1.22 | 0.445 |
|  | 11–20 years | 1.03 | 0.81–1.30 | 0.836 |
|  | 21–30 years | 0.85 | 0.66–1.08 | 0.185 |
|  | 31–40 years | 1.12 | 0.85–1.47 | 0.420 |
| Sex (ref.: Male) | Female | 1.01 | 0.87–1.17 | 0.888 |
| Marital status (ref.: Married) | Single | 1.00 | 0.85–1.16 | 0.969 |
|  | Widowed | 1.19 | 0.54–2.61 | 0.674 |
|  | Divorced | 0.69 | 0.24–2.00 | 0.498 |
| Education (ref.: No schooling) | ★ Primary | 1.29 | 1.06–1.57 | 0.011 |
|  | Secondary | 1.00 | 0.81–1.22 | 0.980 |
|  | Higher/University | 1.11 | 0.60–2.05 | 0.750 |
| Occupation (ref.: Housewife) | Farmer | 1.05 | 0.87–1.28 | 0.556 |
|  | Student/Pupil | 1.18 | 0.98–1.42 | 0.084 |
|  | Teacher/Manager | 1.19 | 0.73–1.94 | 0.477 |
|  | No occupation | 1.19 | 0.82–1.70 | 0.356 |
| Urine (ref.: Normal) | Abnormal (Turbid + Haematic) | 1.13 | 0.97–1.32 | 0.116 |

★ Only factor independently associated with positivity after adjustment ( $p < 0.05$ ). McFadden pseudo- $R^2 = 0.005$ .

## DISCUSSION

### 4.1. Overall Prevalence and Comparison with Other Studies

The overall prevalence of 19.5% (95% CI: 18.4%–20.7%) confirms the significant endemicity of the Ngouri district and is consistent with data available for the Lake Chad basin, although important geographical variations have been documented. In the Torrock sub-prefecture, a study conducted among 1,875 children aged 1–14 years using the same Dawa Mobile Health system reported a prevalence of 24.9% [16]. This slightly higher value is explained by the exclusive focus on children — the most exposed group — and the absence of prior intervention. Further north, around the Ounianga lakes in the Ennedi desert, an exploratory study conducted in 2019 found prevalences as high as 35.3% in Ounianga Kebir and 54.9% in Ounianga Serir [17]. In the Salamat region, a mobile chemoprevention campaign reported a 55% prevalence among school-aged children [18], reflecting a hyperendemic zone with no prior intervention.

Our lower figure most likely reflects the partial impact of a 15-month active intervention, consistent with the initial pre-intervention estimate of 33.1% recorded in the same area. It should be emphasized that our estimate represents a cumulative prevalence in an actively screened population under intervention and cannot be directly compared to population-based prevalence measured outside an intervention context.

### 4.2. Age and Sex Distribution

The absence of statistically significant differences in prevalence by age group (p = 0.163) and sex (p = 0.939) contrasts with most published data from sub-Saharan Africa. A meta-analysis by Ayabina. et al., covering 123 studies across Africa in 2021 found an overall higher prevalence of S. haematobium in males, although only 34% of the sex-related differences observed reached statistical significance, suggesting that sexual dimorphism in infection distribution is far from universal [19]. In Tanzania, after 15 years of mass drug administration, children over 10 years of age remained more frequently infected due to their active lifestyles and frequent contact with contaminated water bodies [20].

In our agro-pastoral Lake Chad context, the homogeneous distribution across sexes and age groups is most plausibly explained by diffuse community-level exposure: the entire population — children, adults, men, and women — shares the same water sources for domestic, agricultural, and pastoral needs. This finding has a direct implication for control strategy: it argues in favor of mass drug administration targeting the entire community, rather than restricting chemoprevention to school-aged children alone.

### 4.3. Primary Education as an Associated Factor

The association between primary education and test positivity (adjusted OR = 1.29; 95% CI: 1.06–1.57; p = 0.011) is the only significant finding from the multivariable model and warrants careful interpretation. In a Nigerian study involving 5,514 primary school children, paternal primary education was significantly associated with infection (adjusted OR = 1.63; 95% CI: 1.01–2.45) [21]. In our setting, this association most likely reflects the behavioral dynamics of children attending primary school (typically aged 6–14 years), who regularly cross wetlands and play near ponds and watercourses on their way to and from school. It is therefore the mobility associated with primary schooling — rather than education level per se — that constitutes the true exposure driver. This interpretation is consistent with the finding that non-schooled subjects, who are generally older, may have developed partial acquired immunity following years of repeated exposure.

### 4.4. Limitations

Several limitations should be considered when interpreting these results. First, the retrospective cross-sectional design does not allow causal inference between the identified factors and infection status. Second, the measured prevalence is cumulative over 15 months in an active intervention zone and does not reflect the instantaneous prevalence in the general population. Third, selection biases cannot be excluded, as participation relied on voluntary demand or community health agent initiative. Foutth, the aggregation of occupational categories into broad groups may have masked finer associations with specific high-risk activities such as fishing or irrigation. Finally, the absence of data on infection intensity and prior praziquantel treatment history limits the epidemiological interpretation of the findings.

## CONCLUSION

This study documents a high prevalence of urogenital schistosomiasis in the Ngouri district, with nearly one in five participants testing positive for Schistosoma haematobium (19.5%; 95% CI: 18.4%–20.7%) after correction of biological inconsistencies. The homogeneous distribution of infection across all demographic subgroups — age, sex, marital status, and occupation — suggests widespread community exposure rather than clearly individualized risk groups. The only factor independently associated with positivity after adjustment was primary education level (adjusted OR = 1.29; p = 0.011), most likely related to the mobility and aquatic behaviors of school-aged children in this agro-pastoral setting.

These findings call for:

- Broad-spectrum preventive chemotherapy targeting the entire community, not limited to school-aged children alone;
- Strengthening of water, sanitation, and hygiene (WASH) infrastructure, particularly around shared water points;
- Consolidation of the Dawa Mobile Health surveillance system, whose feasibility and effectiveness have been demonstrated in this context;
- Systematic data validation at the point of collection to prevent biological inconsistencies such as those identified in this study.

Complementary studies incorporating infection intensity, treatment history, and detailed behavioral data on water contact would be necessary to further refine the understanding of transmission determinants in the Ngouri district.

## Data Availability

All data produced in the present study are available upon reasonable request to the authors

